# MAVS expression in alveolar macrophages is essential for host resistance against *Aspergillus fumigatus*

**DOI:** 10.1101/2021.08.07.21261611

**Authors:** Xi Wang, Cristina Cunha, Madeleine S. Grau, Shelly J. Robertson, João F. Lacerda, António Campos, Katrien Lagrou, Johan Maertens, Sonja M. Best, Agostinho Carvalho, Joshua J. Obar

## Abstract

Our recent data demonstrates a critical role of the RIG-I-like receptor (RLR) family in regulating antifungal immunity against *Aspergillus fumigatus* in a murine model. However, the importance of this pathway in humans and the cell type(s) which utilize this innate immune receptor to detect *A. fumigatus* remains unresolved. Here using patients who underwent hematopoietic stem cell transplantation (HSCT), we demonstrate that a polymorphism in human *MAVS* present in the donor genome was associated with the incidence of invasive pulmonary aspergillosis (IPA). Moreover, in a separate cohort of confirmed IPA patients, polymorphisms in the *IFIH1* gene alter the inflammatory response, including interferon-responsive chemokines. Returning to our murine model, we now demonstrate that CD11c^+^ SiglecF^+^ alveolar macrophages require *Mavs* expression to maintain host resistance against *A. fumigatus*. Our data support the role of MAVS signaling in mediating antifungal immunity in both mice and human at least in part through the role of MAVS-dependent signaling in alveolar macrophages.

## INTRODUCTION

*Aspergillus fumigatus* is a ubiquitous environmental mold that humans inhale on a daily basis. Individuals with normal immune systems readily clear *A. fumigatus* conidia from their airways without complications. In contrast, immunocompromised individuals are at significantly greater risk of developing invasive pulmonary aspergillosis (IPA), including those receiving chemotherapy treatments for cancer and patients receiving immunosuppressive regimens to prevent GVHD following hematopoietic stem cell or solid organ transplantation [1-5]. However, only a small proportion of immunosuppressed patients develop invasive fungal infections indicating that additional risk factors must exist. Numerous human studies have identified genetic polymorphisms in key antifungal pattern recognition receptors and inflammatory cytokines associated with fungal infections (reviewed in [6-8]). Thus, it is important to understand how these responses are coordinated in response to fungal infection of the lungs.

Recently, both type I and type III interferons have been shown to be essential for host resistance against pulmonary *A. fumigatus* challenge [9]. Induction of the type I and type III interferon response following *A. fumigatus* challenge in the mouse model is dependent on both Dectin 1 (*Clec7a*) and MDA5 (*Ifih1*) [10, 11]. Engagement of Dectin 1 leads to Syk activation which drive IRF5 activation for the production of IFNβ following *Candida albicans* challenge [12]. MDA5 engagement by dsRNA leads to its interaction with MAVS resulting in the recruitment of IKKε and TBK1 that activate NFκB and IRF3 and IRF7, respectively, for the production of early cytokines and type I interferons [13]. Genetic polymorphisms within *CLEC7A* have been associated with increased risk of developing IPA [14-16], while the role for genetic polymorphisms within *IFIH1* or *MAVS* regarding susceptibility to IPA has not been explored to date.

Herein, our data demonstrates that genetic polymorphisms within *IFIH1* and *MAVS* alter the production of interferon-dependent chemokines and risk for HSCT patients in developing IPA, respectively. Interestingly, increased risk for developing IPA was associated with genetic variation within *MAVS* only in the donor/hematopoietic compartment. Using our murine model, we identify alveolar macrophages as a key hematopoietic cell in the induction of the MDA5/MAVS-dependent interferon response which is necessary for host resistance against *A. fumigatus*. Overall, our study reveals a critical role for MDA5/MAVS in host anti-fungal immunity in both mice and humans.

## MATERIALS & METHODS

### Mice and Aspergillus fumigatus challenge model

*Ifih1*^*(-/-)*^ (Jackson Laboratory, Stock #015812), *Mavs*^*(fl/fl)*^ [17], and *Mavs*^*(fl/fl)*^ *x Itgax-Cre* mice [17] were bred in-house at Geisel School of Medicine at Dartmouth. C57BL/6J mice were purchased from Jackson Laboratory. All mice were 8-16 weeks of age at the time of challenge. Both female and male mice were used in these studies with no biological difference between the sexes. Animals were not co-housed to equilibrate their microbiota, which is a limitation of this study.

### Preparation of Aspergillus fumigatus conidia and murine challenge model

*A. fumigatus* CEA10 strains were used for this study. *A. fumigatus* was grown on glucose minimal media (GMM) agar plates for 3 days at 37°C. Conidia were harvested by adding 0.01% Tween 80 to plates and gently scraping conidia from the plates using a cell scraper. Conidia were then filtered through sterile Miracloth, were washed, and resuspended in phosphate buffered saline (PBS), and counted on a hemocytometer.

Mice were challenged with *A. fumigatus* conidia by the intratracheal (i.t.) route. Mice were anesthetized by inhalation of isoflurane; subsequently, mice were challenged i.t. with ∼4 × 10^7^ *A. fumigatus* conidia in a volume of 100 µl PBS. At the indicated time after *A. fumigatus* challenge, mice were euthanized using carbon dioxide. Bronchoalveolar lavage fluid (BALF) was collected by washing the lungs with 2 ml of PBS containing 0.05*M* EDTA. BALF was clarified by centrifugation and stored at -20°C until analysis. After centrifugation, the cellular component of the BALF was resuspended in 200 μl of PBS and total BAL cells were determined by hemocytometer count. BALF cells were subsequently spun onto glass slides using a Cytospin4 cytocentrifuge (Thermo Scientific) and stained with the Hema 3(tm) Stat Pack (Fisher Scientific) stain set for differential counting. For histological analysis lungs were filled with and stored in 10% buffered formalin phosphate for at least 24 hours. Lungs were then embedded in paraffin and sectioned into 5-micron sections. Sections were stained with Grocott-Gomori methenamine silver (GMS) using standard histological techniques to assess lung inflammatory infiltrates and fungal germination, respectively. Representative pictures of lung sections were taken using an Olympus BX50WI microscope with a QImaging Retiga 2000R camera.

### Alveolar macrophage isolation and adoptive transfer

Lungs from naïve *Ifih1*^*(-/-)*^, *Mavs*^*(fl/fl)*^ and *Mavs*^*(fl/fl)*^ *x Itgax-Cre* were perfused with 20-30 ml of PBS. Lungs were then removed, injected with 700 μl of collagenase buffer [10ml RPMI, 1ml FBS, 25 μl 0.5*M* CaCl2, 25 μl 0.5*M* MgCl2, 50 μl HEPES, 100 μl L-glutamine, 100 μl Penn-Strep, 2 μl gentamycin, 25 μl DNase (VWR, Catalog # 77001-900), and 2 ml Liberase (Sigma-Aldrich)], and finally placed in 15 ml conical tube containing 2 ml of collagenase buffer. Lungs were digested or 30min at 37°C while shaking at 160-180 rpm. After which, 5 ml of ice cold stop buffer [25 ml RPMI, 1.25 ml FBS, 50 μl 0.5*M* EDTA] was added to each tube. Lungs were pushed through a 70 μm filter to generate a single cell suspension. Single cell lung suspensions were labeled with anti-Siglec-F MicroBeads (Miltenyi Biotec, Cat. No. 130-118-513) and selected for using LS Columns (Miltenyi Biotec, Cat. No. 130-042-401). Siglec F^+^ were eluted from the column, spun down, and resuspended in DMEM at 5×10^6^ cells per ml. Subsequently 5×10^5^ Siglec F^+^ alveolar macrophages were transferred i.t. to *Mavs*^*(fl/fl)*^ *x Itgax-Cre*. Twenty-four hours later mice were challenge with *A. fumigatus* as described above.

### Human HSCT cohort for SNP analysis

A total of 460 hematologic patients undergoing allogeneic hematopoietic stem cell transplantation (HSCT) at the Hospital of Santa Maria, Lisbon and Instituto Português de Oncologia (IPO), Porto, between 2009 and 2015 were included in the study. Cases of IPA were identified and classified as ‘probable’ or ‘proven’ according to the 2008 criteria from the European Organization for Research and Treatment of Cancer/Mycology Study Group (EORTC/MSG) [18]. Exclusion criteria included diagnosis of ‘possible’ IPA, infection with invasive molds other than *Aspergillus* spp. or history of pre-transplant mold infection. In accordance with institutional policies, all patients and donors (or their legal representative) provided informed written consent for data collection, DNA and cell storage, and their use for diagnostic and/or research purposes at the time of referral to transplantation. Study approval was obtained from the institutional review boards (SECVS-125/2014, HSM-632/14 and CES.26/015) and from the National Data Protection Commission (CNPD, 1950/2015) and was in compliance with all local relevant ethical regulations.

Genomic DNA was isolated from whole blood of patients using the QIAcube automated system (Qiagen). SNPs were selected based on their previous association with infection or with putative functional consequences to the gene [19-21]. Genotyping was performed using KASPar assays (LGC Genomics) in an Applied Biosystems 7500 Fast Real-Time PCR system (Thermo Fisher Scientific), according to the manufacturer’s instructions.

### Cytokine analysis of BALF from human IPA patients

BALF and blood samples were collected from hospitalized adult patients (≥18 years of age) at the Leuven University Hospitals, Leuven, Belgium, as previously described [22]. All patients (or their legal representative) provided informed written consent for data collection, DNA and cell storage, and their use for diagnostic and/or research purposes at hospital admission. This study was approved and carried out in accordance with recommendations of the Ethics Subcommittee for Life and Health Sciences of the University of Minho, Portugal (SECVS-125/2014), and the Ethics Committee of the University Hospitals of Leuven, Belgium. Written informed consent was obtained from all subjects in accordance with the Declaration of Helsinki.

For this cytokine analysis, twenty-three cases of “probable” or “proven” IPA were identified according to the standard criteria from the European Organization for Research and Treatment of Cancer/Mycology Study Group (EORTC/MSG) [18] and included for cytokine analysis. Genomic DNA was isolated from EDTA venous blood and genotyped for the *IFIH1* rs1990760 polymorphism. Cytokine levels were determined using the Cytokine & Chemokine 34-Plex Human ProcartaPlex and stratified based on their *IFIH1* rs1990760 genotype.

### Statistical analysis

Statistical significance for *in vitro* and *ex vivo* data was determined by a Mann-Whitney U test, one-way ANOVA using a Bonferroni post-test, or Kruskal-Wallis one-way ANOVA with Dunn’s post-test through the GraphPad Prism 7 software as outlined in the figure legends. Mouse survival data were analyzed with the Mantel-Cox log rank test using GraphPad Prism. For the human HSCT cohort, the probability of IPA resulting from *IFIH1* and *MAVS* SNPs was analyzed using the cumulative incidence method and compared using Gray’s test [23]. The Gray’s test was selected for the analysis because, in contrast with the commonly used Kaplan-Meier and Cox models, it allows accounting for multiple competing risks and provides a better estimation for the risk of the main outcome of interest, in this case the development of IPA [24]. Cumulative incidences were computed with the *cmprsk* package for R version 2.10.1 [25], with censoring of data at the date of last follow-up visit and defining relapse and death as competing events. A period of 24 months after transplant was chosen to include all cases of IPA.

### Study approvals

All animal experiments were approved by the Dartmouth College Institutional Animal Care and Use Committee under protocol number 00002168. For our human studies approval was obtained from the institutional review boards (SECVS-125/2014, HSM-632/14 and CES.26/015) and from the National Data Protection Commission (CNPD, 1950/2015) and was in compliance with all local relevant ethical regulations.

## RESULTS

### Single Nucleotide Polymorphisms (SNPs) in *IFIH1* and *MAVS* influence the risk for developing invasive pulmonary aspergillosis

Our recent mouse data suggests that MDA5/MAVS signaling is critical in maintaining host resistance against pulmonary challenge with *A. fumigatus* [11]. Therefore, we wanted to explore the role of these molecules in a human cohort comprised of 460 HSCT patients (Supplemental Table 1). Missense SNPs within the coding region of *IFIH1* (rs1990760 and rs3747517) are associated with autoimmune conditions, particularly interferonopathies [19]. Moreover, a missense SNP in *MAVS* (rs17857295) is associated with altered type I interferon regulation [21]. While an individual genotype for each SNP was not associated with risk of developing IPA (Supplemental Figure 1), we further examined these SNPs using a recessive genetic model because the autoimmune risk allele in *IFIH1* rs1990760 has been shown to work in a dominant fashion [19]. Notably, we found a significant association between the CC genotype at rs1990760 in *IFIH1* and the risk of developing IPA in HSCT recipients (Figure 1A). This increased risk occurred when the variant was carried by the recipient in a recessive genetic model, but not when it was carried by the donor. In contrast, no association between the *IFIH1* rs3747517 SNP and the risk of developing IPA was found (Figure 1B). Additionally, we also found a significant association between the *MAVS* rs17857295 SNP and the risk of developing IPA in HSCT recipients (Figure 1C). This increased risk occurred when the variant was carried by the donor in a recessive genetic model, but not when it was carried by the recipient. Moreover, the risk from the donor *MAVS* rs17857295 SNP held up to multivariate analysis accounting for relevant clinical risk factors (Table 1).

**Figure 1.**
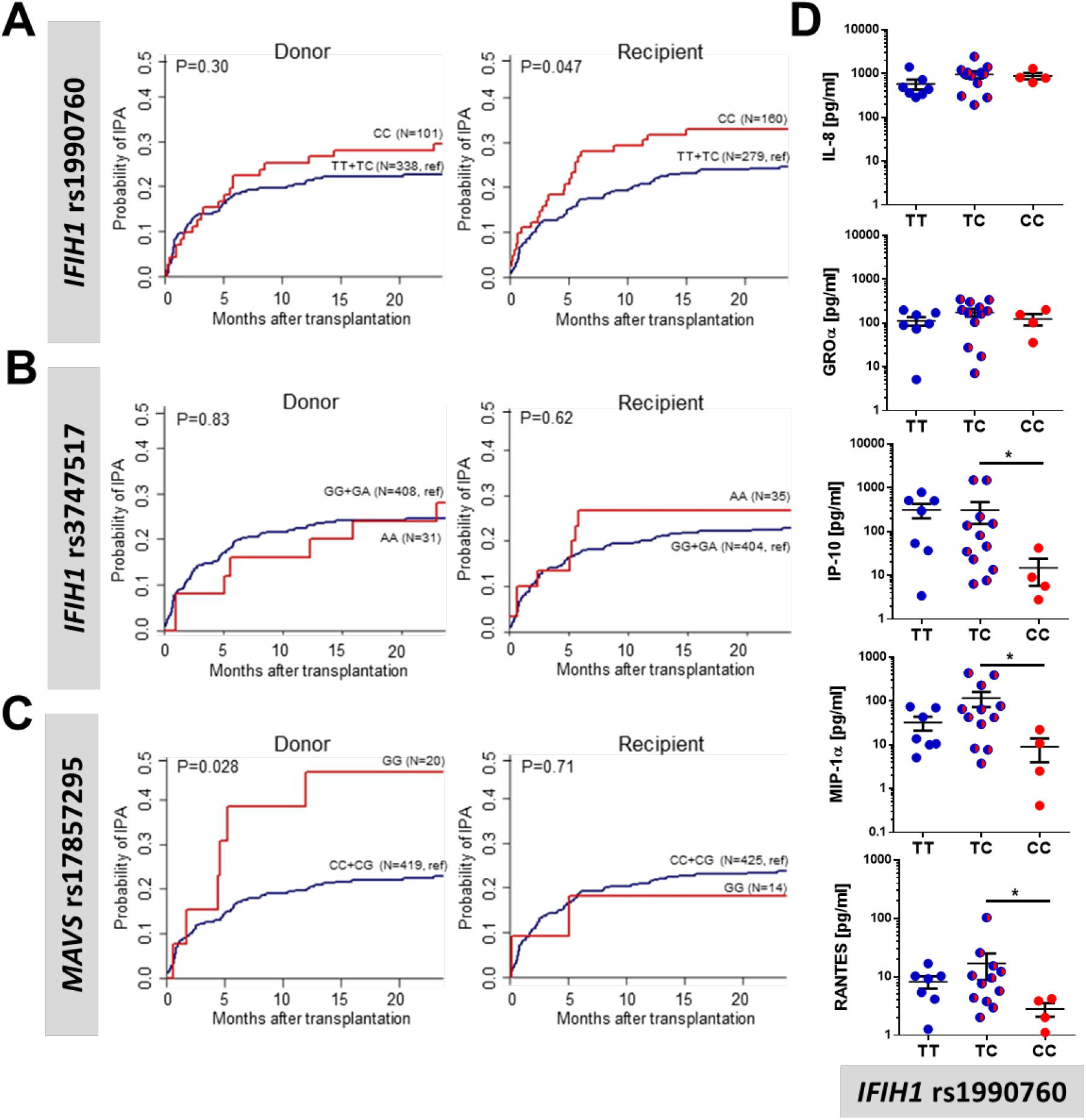
*IFIH1* and *MAVS* polymorphisms are associated with invasive pulmonary aspergillosis in human transplant patients using a recessive allele model. Cumulative incidence analysis of invasive aspergillosis after transplantation according to donor or recipient genotypes at rs1990760 in *IFIH1* **(A)**, rs3747517 in *IFIH1* **(B)**, or rs17857295 in *MAVS* **(C)** over 24 months after HSCT. Data were analyzed by two-sided Gray’s test. **(D)** Inflammatory cytokine levels in the bronchoalveolar lavage fluid from 23 patients with invasive pulmonary aspergillosis were measured using a 32-plex ProCarta Luminex assay and plotted based on their *IFIH1* rs1990760 genotype. Data were analyzed using a Mann-Whitney U-test (* p < 0.05).

**Table 1.** Multivariate analysis of the association of *IFIH1 and MAVS* SNPs with the risk of invasive pulmonary aspergillosis among transplant recipients.

| Genetic/clinical variables | Adjusted HR<sup>†</sup><br>(95% CI) | P value |
| --- | --- | --- |
| Donor rs17857295 in *MAVS* | 2.39 (1.07 – 5.32) | 0.033 |
| Recipient rs1990760 in *IFIH1* | 1.46 (0.89 – 2.40) | 0.121 |
| Acute GVHD grades III-IV | 1.73 (0.94 – 3.19) | 0.047 |

HR, hazard ratio; CI, confidence interval. Multivariate analyses were based on the subdistribution regression model of Fine and Gray. <sup>†</sup>Hazard ratios were adjusted for patient age and gender, and clinical variables with a P<0.15 in the univariate analyses. Only the clinical variables remaining significant after adjustment are shown.

To demonstrate a functional effect of the rs1990760 SNP in *IFIH1* and the associated amino acid substitution on the function of MDA5, we analyzed the inflammatory response in the bronchoalveolar lavage fluid (BALF) of 23 patients with IPA after stratification by genotypes at rs1990760 in *IFIH1* (Supplemental Table 2). We found that individuals with the CC genotype, which is associated with increased risk of developing IPA (Figure 1A), had a significant reduction in IP-10 (CXCL10), RANTES (CCL5), and MIP-1α (CCL3), but not IL-8 and GROα (CXCL1) (Figure 1D). IP-10 (CXCL10), RANTES (CCL5), and MIP-1α (CCL3) are known to be interferon-responsive genes, while IL-8 and GROα (CXCL1) are not [26]. In the BALF samples from this patient cohort, the levels of IFN-α were below the detection limit (data not shown). Given the relatively rare frequency of the rs17857295 genotype in *MAVS*, it was not possible to analyze cytokine production according to *MAVS* genotype in the BAL samples (data not shown). Overall, these data support an important role for MDA5/MAVS signaling in regulating the inflammatory response and host resistance against *Aspergillus* spp. in humans.

### Deletion of *Mavs* in CD11c^+^ cells result in increased susceptibility to *Aspergillus fumigatus* in mice

Given the observation that patients in the HSCT cohort with the *MAVS* rs17857295 polymorphism within the donor population had altered risk for developing IPA, we next wanted to assess which hematopoietic cell population(s) in our invasive aspergillosis murine model relies on *Mavs* to maintain host resistance against *A. fumigatus*. To directly address which cells must express *Mavs* we utilized the recently developed *Mavs*^*(fl/fl)*^ *x Itgax-Cre* conditional knock-out model, hereafter referred to as *Mavs*^*(Cd11c/Cd11c)*^, that will delete *Mavs* in all CD11c-expressing cells [17], including both dendritic cells and alveolar macrophages. To test of role of *Mavs* expression in CD11c-expressing cells for the maintenance of host resistance against *A. fumigatus*, we challenged C57BL/6J, *Mavs*^*(fl/fl)*^, and *Mavs*^*(Cd11c/Cd11c)*^ mice with 4×10^7^ conidia of the CEA10 strain and monitored survival over nine days. *Mavs*^*(Cd11c/Cd11c)*^ mice were more susceptible to pulmonary challenge with *A. fumigatus* than either C57BL/6J or *Mavs*^*(fl/fl)*^ mice (Figure 2A; Mantel-Cox log rank test, p < 0.0001). Mice completely lacking either *Ifih1* or *Mavs* have decreased neutrophil recruitment, which was associated with increased fungal germination after pulmonary challenge with *A. fumigatus* [11]. Thus, we assessed fungal germination in the lung by histological analysis at 48 h after conidial instillation. Strikingly, GMS staining of lung tissue from *Mavs*^*(Cd11c/Cd11c)*^ mice revealed high levels of *A. fumigatus* germination at this time compared with control *Mavs*^*(fl/fl)*^ mice (Figure 2B). When the percentage of germinated *A. fumigatus* was quantified, *Mavs*^*(fl/fl)*^ mice displayed low levels of fungal germination (23.5% ± 9.1) compared with *Mavs*^*(Cd11c/Cd11c)*^ mice (60.1% ± 8.2). Finally, we assessed inflammatory cell accumulation in the airways. The increased susceptibility of *Mavs*^*(Cd11c/Cd11c)*^ mice to *A. fumigatus* challenge was associated with decreased accumulation of neutrophils in the airways compared with *Mavs*^*(fl/fl)*^ mice (Figure 2C). These data match our prior observations with mice completely lacking either *Ifih1* or *Mavs* having increased IPA susceptibility [11], suggesting that *Mavs* expression within CD11c-expressing cells is critical for host resistance against *A. fumigatus* challenge.

**Figure 2.**
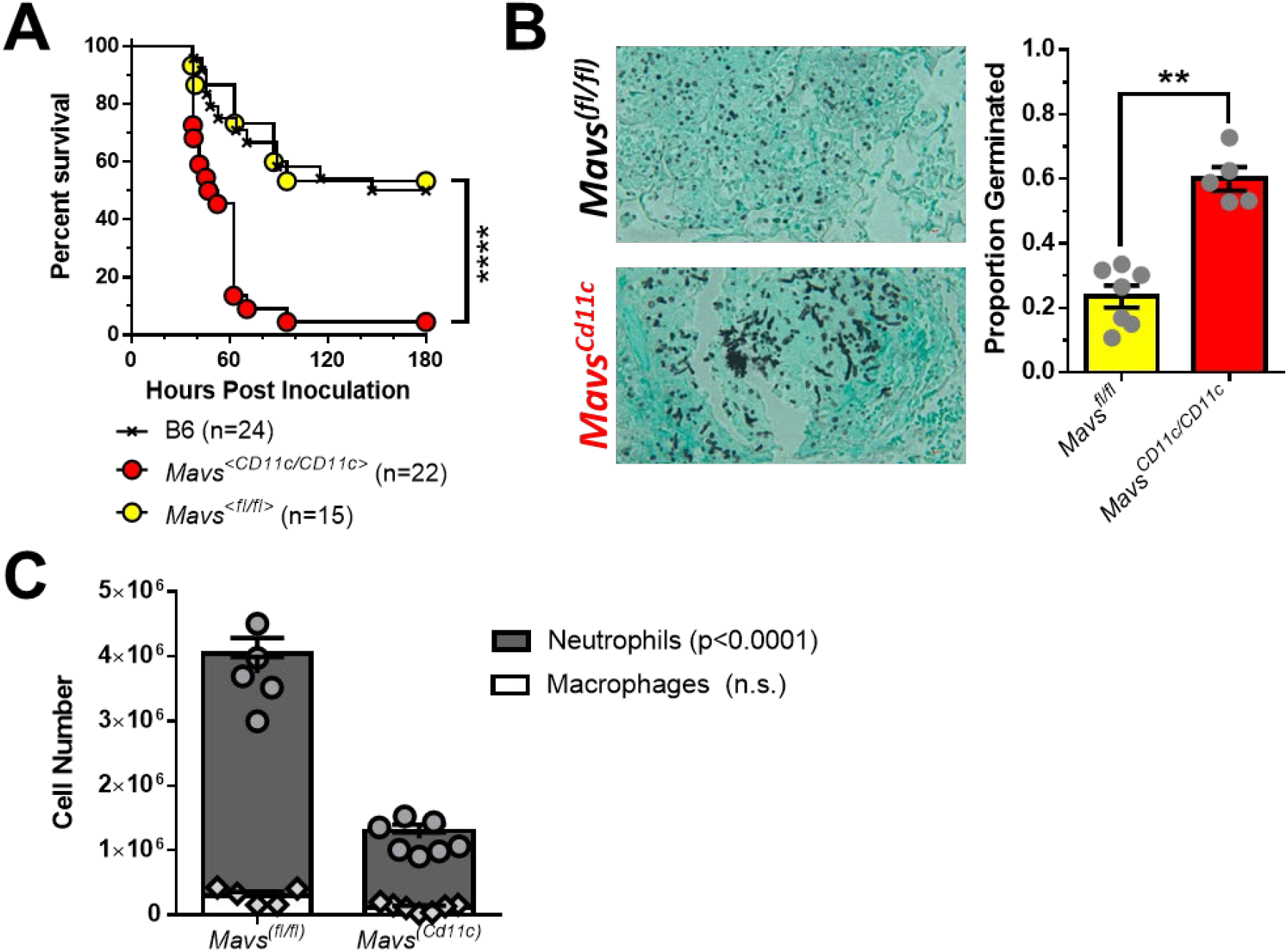
Mavs-dependent responses are essential in CD11c^+^ cells for host resistance against Aspergillus fumigatus. *Mavs* ^*<Cd11c/Cd11c>*^, *Mavs*^*<fl/fl>*^ and C57BL/6J mice were challenged i.t. with 4×10^7^ resting conidia of the CEA10 isolate of *Aspergillus fumigatus*. **(A)** Survival analysis in immune-competent wild-type and knock-out mice were tracked over the first 9 days. ****, P < 0.0001 by Mantel-Cox log rank test. **(B)** Forty hours after *A. fumigatus* challenge mice were euthanized and fungal germination was assessed in the lungs by GMS staining. **, P < 0.01 by Mann-Whitney U-test. **(C)** At the same time point, cell differentials in the lung airways was determined by differential staining of BALF cytospins. Statistical significance was determined using a two-way ANOVA with a Sidak’s post-test.

### Alveolar macrophage transfer to *Mavs*^*(Cd11c/Cd11c)*^ conditional knock-out mice reduces their susceptibility to *Aspergillus fumigatus* in mice

Multiple cells in the lungs can express *Igtax* (CD11c) including alveolar macrophages, CD103^+^ dendritic cells, CD11b^+^ dendritic cells, monocyte-derived dendritic cells, and plasmacytoid dendritic cells, all of which have be implicated in the antifungal immune response against *A. fumigatus* [27-33]. Utilizing the publicly available data assembled by the ImmGen Consortium [34], in the lungs we see that *Mavs* is highly expressed in Siglec F^+^ alveolar macrophages, but also moderately expressed in both monocytes and dendritic cells (data not shown). Interestingly, following RSV infection alveolar macrophages have been shown to be the key cell type for initiating the type I interferon through a MAVS-dependent mechanism [35, 36]. While alveolar macrophages largely populate the lungs during embryogenesis and are maintained by local proliferation during homeostasis [37, 38] following conditioning for HSC transplantation alveolar macrophage can be repopulated from the donor HSC compartment [39-42]. Therefore, to specifically address the role of *Mavs* in alveolar macrophages, we purified Siglec F^+^ cells from the lungs of naïve *Mavs*^*(fl/fl)*^ or *Mavs*^*(Cd11c/Cd11c)*^ mice and transferred 5×10^5^ Siglec F^+^ cells intratracheally into *Mavs*^*(Cd11c/Cd11c)*^ mice one day prior to challenging with 4×10^7^ conidia of the CEA10 strain (Figure 3A). Forty-eight hours after challenge, we examined the lungs by GMS staining of lung tissue from *Mavs*^*(Cd11c/Cd11c)*^ mice receiving the *Mavs*^*(Cd11c/Cd11c)*^ Siglec F^+^ cells and revealed the presence of high levels of germinated *A. fumigatus* (68.1% ± 12.6), while that was not observed to the same extent in *Mavs*^*(Cd11c/Cd11c)*^ mice receiving the *Mavs*^*(fl/fl)*^ Siglec F^+^ cells (31.4% ± 6.7) (Figure 3B). This is in line with control *Mavs*^*(Cd11c/Cd11c)*^ mice or *Mavs*^*(fl/fl)*^ mice receiving DMEM (vehicle) administered intratracheally. When we examined the accumulation of leukocytes in the airways by differential cytospin analysis we found that *Mavs*^*(Cd11c/Cd11c)*^ mice receiving *Mavs*^*(Cd11c/Cd11c)*^ Siglec F^+^ cells had reduced numbers of neutrophils when compared with *Mavs*^*(Cd11c/Cd11c)*^ mice receiving *Mavs*^*(fl/fl)*^ Siglec F^+^ cells (Figure 3C). Strikingly, when we examined macrophage numbers in the airways, we found *Mavs*^*(Cd11c/Cd11c)*^ mice receiving *Mavs*^*(fl/fl)*^ Siglec F^+^ cells had many macrophages present, but *Mavs*^*(Cd11c/Cd11c)*^ mice receiving *Mavs*^*(Cd11c/Cd11c)*^ Siglec F^+^ cells had few macrophages in their airways at 48 hours post-inoculation with *A. fumigatus* (Figure 3D). Importantly, the decrease in macrophages in the airways is not observed in naïve *Mavs*^*(Cd11c/Cd11c)*^ mice (Supplemental Figure 2A). Additionally, *Mavs*^*(Cd11c/Cd11c)*^ mice receiving an i.t. transfer of *Mavs*^*(Cd11c/Cd11c)*^ Siglec F^+^ cells 24h prior only had a moderate decrease in alveolar macrophage numbers (Supplemental Figure 2B).

**Figure 3.**
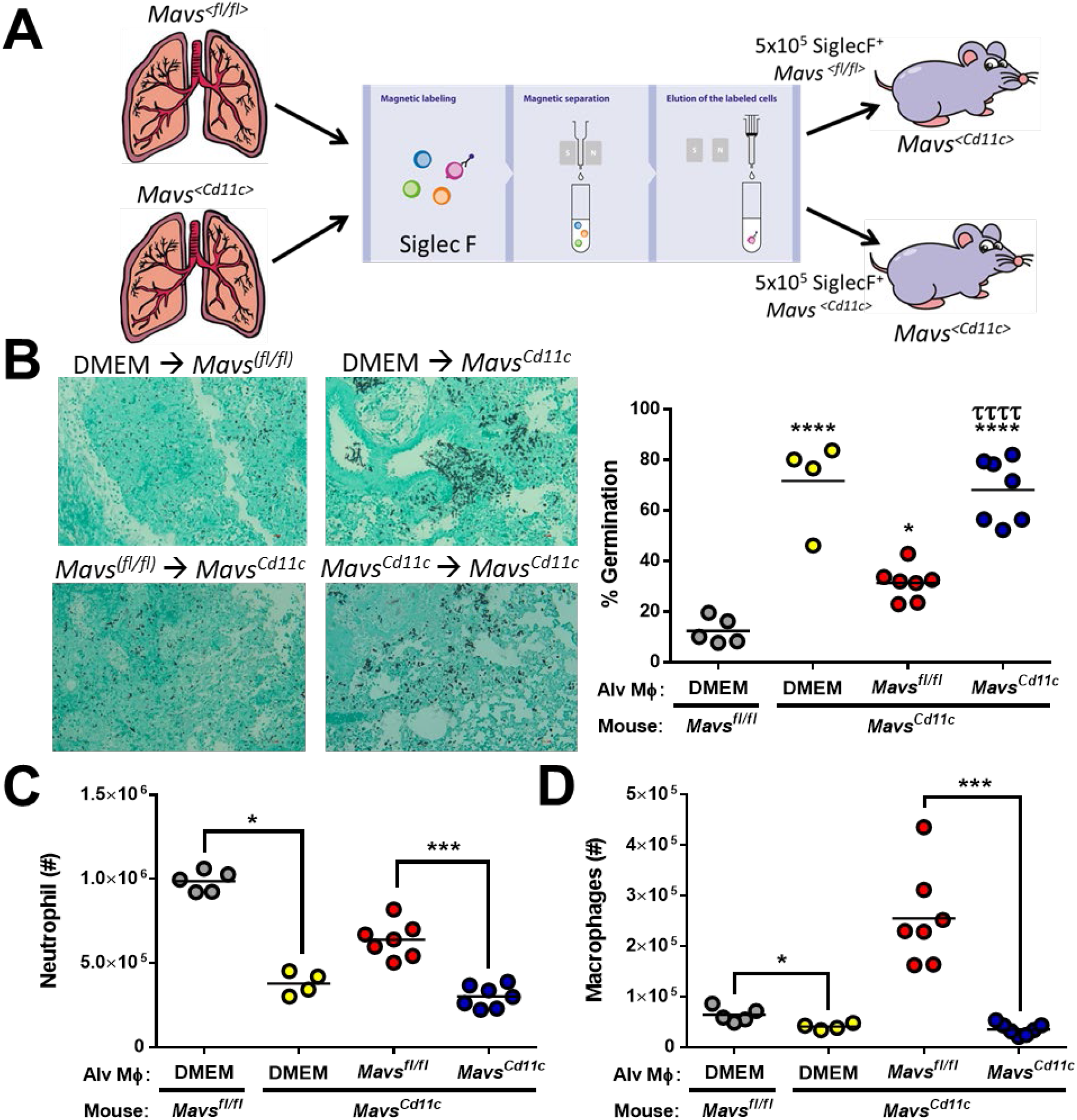
Alveolar macrophages require a Mavs-dependent response to maintain host resistance against Aspergillus fumigatus. **(A)** *Mavs*^*<Cd11c/Cd11c>*^ mice were complemented with 5×10^5^ *Mavs*^*<fl/fl>*^ or *Mavs* ^*<Cd11c/Cd11c>*^ alveolar macrophages (Siglec F^+^) given intranasally. One day later mice were challenged i.t. with 4×10^7^ resting conidia of the CEA10 isolate of *Aspergillus fumigatus*. Forty hours after *A. fumigatus* challenge mice were euthanized. **(B** Fungal germination was assessed in the lungs by GMS staining. Representative 20x GMS images are shown. Statistically significant different were determined using a one-way ANOVA with a Tukey’s post-test: *p<0.05 (vs. DMEM → *Mavs*^*(fl//fl)*^), ****p<0.0001 (vs. DMEM → *Mavs*^*(fl//fl)*^), and ττττ p<0.0005 (vs. *Mavs*^*(fl//fl)*^→ *Mavs*^*(Cd11c)*^). Neutrophils **(C)** and macrophages **(D)** in the lung airways was determined by differential staining of BALF cytospins.

Next, to examine the role of *Ifih1* in alveolar macrophages, we purified Siglec F^+^ cells from the lungs of naïve *Mavs*^*(fl/fl)*^ or *Ifih1*^*(-/-)*^ mice and transferred 5×10^5^ Siglec F^+^ cells intratracheally into *Mavs*^*(Cd11c/Cd11c)*^ mice one day prior to challenging with 4×10^7^ conidia of the CEA10 strain (Supplemental Figure 3). Forty-eight hours after challenge, we examined the lungs by GMS staining of lung tissue from *Mavs*^*(Cd11c/Cd11c)*^ mice receiving either DMEM or *Ifih1*^*(-/-)*^ Siglec F^+^ cells, revealing the presence of high levels of germinated *A. fumigatus*, which was not observed to the same extent in *Mavs*^*(Cd11c/Cd11c)*^ mice receiving the *Mavs*^*(fl/fl)*^ Siglec F^+^ cells (Supplemental Figure 3). Taken all together, these data demonstrate that *Ifih1* and *Mavs* expression within Siglec F^+^ alveolar macrophages is critical for host resistance against *A. fumigatus*.

## DISCUSSION

Humans inhale *Aspergillus fumigatus* spores on a daily basis, but individuals with healthy immune systems readily clear *A. fumigatus* conidia from their airways without problems. One lung sentinel cell that can be critical in the clearance of *A. fumigatus* is the alveolar macrophage. Alveolar macrophages are known to phagocytose and destroy *A. fumigatus* [31], but their role in host resistance has remained controversial [33, 43]. Patients undergoing HSCT are at increased risk of developing IPA [1-5]. In early studies from patients undergoing HSCT, alveolar macrophages are dysfunctional [41] and found at decreased in numbers [40-42] for at least 50 days post-transplantation. However, we currently do not understand the effects of modern HSCT conditioning regiments on alveolar macrophages function and numbers, which is a limitation of our study. Either way it is important to understand the role of alveolar macrophages in the initial inflammatory response induced by *A. fumigatus*. These lung sentinel alveolar macrophages are also known to participate in the inflammatory response induced by *A. fumigatus* [44], but their role in driving the interferon response following *A. fumigatus* challenge was not addressed. During respiratory syncytial virus infection alveolar macrophages have been demonstrated to be key drivers of the type I interferon response through a MAVS-dependent mechanism [35, 36]. We recently describe a critical role for MDA5/MAVS-dependent induction of interferons following *A. fumigatus* challenge [11], but the cellular localization of MAVS-dependent signaling was not elucidated. Here we demonstrated that *Mavs* expression in alveolar macrophages was at least partially necessary for host resistance against *A. fumigatus*, through the regulation of neutrophil accumulation in the lung for the prevention of fungal growth. While alveolar macrophage transfer to *Mavs*^*(Cd11c/Cd11c)*^ mice largely restores host resistance, it was not absolute suggesting additional CD11c^+^ cells might require *Mavs* expression for resistance. One potential candidate would be plasmacytoid dendritic cells that are known to produce type I interferons after fungal challenge [27] and are essential for host resistance *A. fumigatus* [45], but the importance of MAVS signaling in plasmacytoid dendritic cells in driving the interferon response remains controversial. Additionally, in a dendritic cell vaccination model, *A. fumigatus* conidial RNA was shown to be sufficient for the activation of dendritic cells to preferentially prime a Th1 immune response [46], which was at least partially dependent on TLR3 [47]. The role of MAVS signaling in other CD11c^+^ cell populations will be explored in future studies. Alveolar macrophages also participate in the inflammatory response to other fungal pathogens including *Cryptococcus neoformans* [48], *Pneumocystis* spp. [49], and *Rhizopus* spp. [50]. Interestingly, heterogeneity within the alveolar macrophage population has been demonstrated particularly in regard to CXCL2 expression [48], which could be important in our findings showing that the mice lacking *Ifih1* or *Mavs* have decreased neutrophil accumulation following *A. fumigatus* challenge [11]. Additionally, Jakubzick and colleagues have identified significant heterogeneity within human alveolar macrophages using single cell RNA-Seq, which include the identification of novel IFN responsive alveolar macrophage populations and a number of chemokine producing populations [51]. Understanding the specific role of these alveolar macrophage populations in host resistance is the goal of future studies.

In addition to the role of alveolar macrophages in the interferon-dependent antifungal immune response that we describe herein, it has previously been shown that *in vitro* human bronchial epithelial cells can secrete IFNβ and CXCL10 in response *A. fumigatus* spore challenge [52]. Following respiratory virus infections, the lung epithelium can be a critical source of antiviral interferons, which can be driven by both TLRs and RLRs [53, 54]. In the human bronchial epithelial cell model, Bals and colleagues demonstrated that fungal dsRNA which was transfected into the human bronchial epithelial cells was sufficient for inducing IFNβ and CXCL10 secretion [52], which is similar to our previous findings using in murine fibroblast [11]. However, while Bals and colleagues demonstrated that *A. fumigatus* dsRNA was sufficient for the activation of TLR3-TRIF signaling, they did not examine the necessity of this pathway in driving IFNβ and CXCL10 secretion by the human bronchial epithelial cells [52]. Interestingly, in our preliminary studies, cytosolic delivery of *A. fumigatus* dsRNA drives both MDA5/MAVS-dependent and TRIF-dependent IFNα and CXCL10 response in murine fibroblasts (data not shown). Moreover, Romani and colleagues have found both *Tlr3-* and *Ticam1* (TRIF)-deficient mice are highly susceptible to pulmonary *A. fumigatus* challenge [47, 55], and this was due entirely to TRIF-dependent response in epithelial cells [55]. Interestingly, in our HSCT patients, the *IFIH1* SNP association was found in the host genotype. Taken together, these data suggest the lung epithelial cells could be another potential source of IFNs after *A. fumigatus* challenge which needs to be explored in future studies.

While our previous studies have shown the importance of MDA5/MAVS signaling [11] and type I/III interferons [9] in host resistance against *A. fumigatus* in the murine model, the importance of these pathways in humans has not been described. Herein, we have found that antifungal role of MDA5/MAVS signaling is also critical in humans. Our data from a cohort of HSCT patients demonstrate that SNPs in *IFIH* (rs1990760) and *MAVS* (rs17857295) are associated with an altered risk of developing IPA. Interestingly, SNPs within *IFIH1* have been associated with numerous autoimmune diseases, particularly those that are highly dependent on type I interferons for their progression [19, 56]. Recent evidence suggests that the autoimmune risk genotype of the rs1990760 SNP in *IFIH1* (TT) has functional consequences in human PBMCs, specifically it is associated with elevated basal levels of *IFNB1* [19]. Knock-in of the *IFIH1* rs1990760 autoimmune risk genotype (TT) into mice not only recapitulates the susceptibility to autoimmune diseases, but also results in increased host resistance against encephalomyocarditis virus (ECMV), a picornavirus specifically recognized by MDA5 [57], and elevated *Ifnb1* expression [19]. Moreover, in human pancreatic islets it has been shown that the *IFIH1* rs1990760 TC genotype also drove enhanced interferon signaling after Coxsackievirus infection [20]. This enhanced interferon response during Coxsackievirus infection was associated with greater signaling from the peroxisomes by the *IFIH1* rs1990760 TC genotype [20], which has previously been shown to preferentially induce a type III interferon response [58, 59]. Interestingly, our SNP analysis of a human HSCT cohort found that the T allele of *IFIH1* rs1990760, which as just mentioned is associated with an increased interferon response [19, 20], was associated with a reduced risk for the development of IPA. In addition to the *IFIH1* rs1990760 SNP, we also found a clinical association with the rs17857295 SNP in *MAVS* with the risk of developing IPA after HSCT. Much less is known about the functional outcome of the rs17857295 SNP in *MAVS*, but one report suggests overexpression of the GG genotype in 293T cells stably knocked-down for the endogenous *MAVS* allele resulted in an inability to induce *Ifnb1* expression following PolyI:C stimulation [21]. Overall, our analysis suggests that HSCT patients harboring these SNPs display a reduced ability to trigger an interferon response and are at greater risk of developing IPA.

In contrast to our clinical data from HSCT patients who develop IPA, the T allele at rs1990760 in *IFIH1* is associated with an increased risk for the development of chronic mucocutaneous candidiasis [60]. Again, the T allele at rs1990760 is associated with an increased interferon response [19, 20], but the lack of IFNAR-dependent signaling leads to increased susceptibility to invasive candidiasis in mice [12]. In their study Netea and colleagues did not observe altered *Ifnb* expression in the absence of MDA5 after stimulation with *Candida albicans* hyphae [60], but a role for MDA5 in regulating other interferons was not explored. Our data with *A. fumigatus* suggests that MDA5/MAVS-signaling are more important in the regulation of type III interferons (IFNλ/IL-28), rather than type I interferons (IFNα/β). Excessive inflammation and tissue pathology can be critical in mediating disease during invasive candidiasis [61]. Rivera and colleagues demonstrated that type III interferon (IFNλ/IL-28) signaling enhances the production of reactive oxygen species by neutrophils following *A. fumigatus* [9], which could enhance disease during invasive candidiasis. Thus, there appears to be an interesting dichotomy in MDA5 signaling that is crucial for tuning host resistance against different fungal pathogens and warrants further research.

## Data Availability

All data will be available upon request

## AUTHOR CONTRIBUTIONS

Conceived and designed the experiments: XW, AC, JJO. Performed the experiments: XW, CC, MG. Analyzed the data: XW, CC, MG, AC, JJO. Provided clinical samples: JFL, AC Jr, KL, JM. Wrote the paper: XW, JJO.

## ACKNOWLEDGEMENTS

Thank you to Drs. Robert Cramer, Claudia Jakubzick, and David Leib (Geisel School of Medicine at Dartmouth) for helpful discussion on this project.

## FOOTNOTES

Research in this study was supported in part by institutional startup funds to JJO in part through the Dartmouth Lung Biology Center for Molecular, Cellular, and Translational Research grant P30 GM106394 (PI: Bruce A. Stanton) and Center for Molecular, Cellular and Translational Immunological Research grant P30 GM103415 (PI: William R. Green). JJO was partially supported by a Munck-Pfefferkorn Award from Dartmouth College and NIH R01 AI139133 grant. SMB is supported by the Division of Intramural Research, National Institutes of Health, National Institute of Allergy and Infectious Diseases. AC and CC were supported by the Fundação para a Ciência e a Tecnologia (FCT) (PTDC/SAU-SER/29635/2017, PTDC/MED-GEN/28778/2017, UIDB/50026/2020, UIDP/50026/2020, and CEECIND/04058/2018), the Northern Portugal Regional Operational Programme (NORTE 2020), under the Portugal 2020 Partnership Agreement, through the European Regional Development Fund (ERDF) (NORTE-01-0145-FEDER-000039), the European Union’s Horizon 2020 research and innovation programme under grant agreement no. 847507, and the “la Caixa” Foundation (ID 100010434) and FCT under the agreement LCF/PR/HR17/52190003. The funders had no role in the preparation or publication of the manuscript.

## Abbrevations

IPA: invasive pulmonary aspergillosis
HCST: hematopoietic stem cell transplantation
GMM: glucose minimal media
PBS: phosphate buffered saline
i.t.: intratracheal
BALF: bronchoalveolar lavage fluid
GMS: Grocott-Gomori methenamine silver
SNP: single nucleotide polymorphism

---

**Supplemental Table 1.** Baseline characteristics of transplant recipients enrolled in the genetic association study.

| Variables | IPA<br>(n=91) | No IPA<br>(n=348) | P value |
| --- | --- | --- | --- |
| **Age at transplantation, no (%)** |  |  |  |
| ≤20 years | 13 (14.3) | 69 (19.8) | 0.264 |
| 21 – 40 years | 23 (25.3) | 101 (29.0) |  |
| >40 years | 55 (60.4) | 178 (51.2) |  |
| **Gender, no (%)** |  |  |  |
| Female | 38 (41.8) | 150 (43.1) | 0.859 |
| Male | 53 (58.2) | 198 (56.9) |  |
| **Underlying disease, no. (%)** |  |  |  |
| Acute leukemia | 49 (53.8) | 179 (51.5) | 0.115 |
| Chronic lymphoproliferative diseases | 14 (15.4) | 69 (19.8) |  |
| Myelodysplastic/myeloproliferative diseases | 13 (14.3) | 30 (8.6) |  |
| Chronic myeloproliferative diseases | 7 (7.7) | 20 (5.7) |  |
| Aplastic anemia | 6 (6.6) | 17 (4.9) |  |
| Other | 2 (2.2) | 33 (9.5) |  |
| **Transplantation type, no. (%)** |  |  |  |
| Matched, related | 34 (37.4) | 169 (48.6) | 0.037 |
| Matched, unrelated | 33 (36.3) | 81 (23.3) |  |
| Mismatched, related | 0 (0.0) | 7 (2.0) |  |
| Mismatched, unrelated | 24 (26.4) | 91 (26.2) |  |
| **Graft source, no. (%)** |  |  |  |
| Peripheral blood | 80 (87.9) | 287 (82.5) | 0.506 |
| Bone-marrow | 10 (11.0) | 53 (15.2) |  |
| Cord blood | 1 (1.1) | 8 (2.3) |  |
| **Disease stage, no. (%)** |  |  |  |
| First complete remission | 49 (53.8) | 188 (54.0) | 0.800 |
| Second or subsequent remission, or relapse | 13 (14.3) | 59 (17.0) |  |
| Active disease | 29 (31.9) | 101 (29.0) |  |
| **Conditioning regimen, no (%)** |  |  |  |
| RIC | 68 (74.7) | 228 (65.5) | 0.091 |
| Myeloablative | 23 (25.3) | 120 (34.5) |  |
| **CMV serostatus of donor and recipient, no. (%)** |  |  |  |
| D-/R+ or D+/R+ | 80 (87.9) | 313 (89.9) | 0.504 |
| D-/R- or D+/R- | 11 (12.1) | 35 (10.1) |  |
| **Duration of neutropenia, mean days (range)<sup>†</sup>** | 13.1 (8 – 39) | 13.5 (5 – 35) | 0.460 |
| **Acute GVHD, no. (%)** |  |  |  |
| No GVHD or grades I – II | 63 (69.2) | 302 (86.8) | 0.0002 |
| Grades III – IV | 28 (30.8) | 46 (13.2) |  |
| **Antifungal prophylaxis, no. (%)<sup>‡</sup>** |  |  |  |
| Fluconazole | 42 (46.2) | 117 (33.6) | 0.002 |
| Posaconazole | 26 (28.6) | 107 (30.8) |  |
| Other | 9 (9.9) | 14 (4.0) |  |
| None or unknown | 14 (15.4) | 110 (31.6) |  |

Twenty-one patients with “possible” IPA were excluded. Chronic lymphoproliferative diseases included cases of chronic lymphocytic leukemia, multiple myeloma, and B- and T-cell lymphomas. Chronic myeloproliferative diseases included cases of chronic myelogenous leukemia and primary myelofibrosis. Other diseases included cases of idiopathic medullar aplasia, lymphohistiocytosis, hemoglobinopathies and paroxysmal nocturnal hemoglobinuria. RIC, reduced intensity conditioning; CMV, cytomegalovirus; D, donor; R, recipient; GVHD, graft-versus-host-disease. <sup>†</sup>Neutropenia was defined as $\leq 0.5 \times 10^9$ cells/L. <sup>‡</sup>Other antifungals used in prophylaxis included voriconazole, liposomal amphotericin B, itraconazole and caspofungin. P values were calculated by Fisher’s exact probability t-test or Student’s t-test for continuous variables.

**Supplemental Table 2.** Demographics and clinical characteristics of the patients used for BAL cytokine analysis.

| **Variables** | **IPA**<br>**(n=23)** |
| --- | --- |
| **Age, mean ± SD** | 60.9 ± 9.0 |
| **Male, no (%)** | 15 (65.0) |
| **Underlying disease, no (%) \*** |  |
| Acute leukemia | 8 (34.8) |
| Allogeneic HSCT | 5 (21.7) |
| Chronic lymphoproliferative diseases | 4 (17.4) |
| SOT | 3 (13.0) |
| Influenza A (H1N1) | 2 (8.7) |
| Lung cancer | 1 (4.3) |
| **ICU admission** | 11 (47.8) |
| **Mechanical ventilation** | 8 (34.8) |
| **Severe neutropenia, no (%) †** | 11 (47.8) |
| **BAL cell counts, mean (range) ‡** |  |
| Neutrophils | 2.9 (0.0–12.6) |
| Lymphocytes | 0.6 (0.0–53.0) |
| **Immunosuppression, no (%)** |  |
| Steroids | 10 (43.5) |
| Other immunosuppressive regimens | 3 (13.0) |
| None | 10 (43.5) |

IPA, invasive pulmonary aspergillosis; HSCT, hematopoietic stem-cell transplantation; SOT, solid organ transplantation; ICU, intensive care unit; BAL, bronchoalveolar lavage; \*SOT included lung, kidney, and liver transplants. †Severe neutropenia was defined as $\leq 0.5 \times 10^9$ cells/L. ‡Cell counts in BAL were expressed as the number $\times 10^3$ cells/ $\mu$ L. P values were calculated by Fisher's exact probability t-test for categorical variables, or by Student's t-test or Mann-Whitney U test for continuous variables.

**Supplemental Figure 1.**
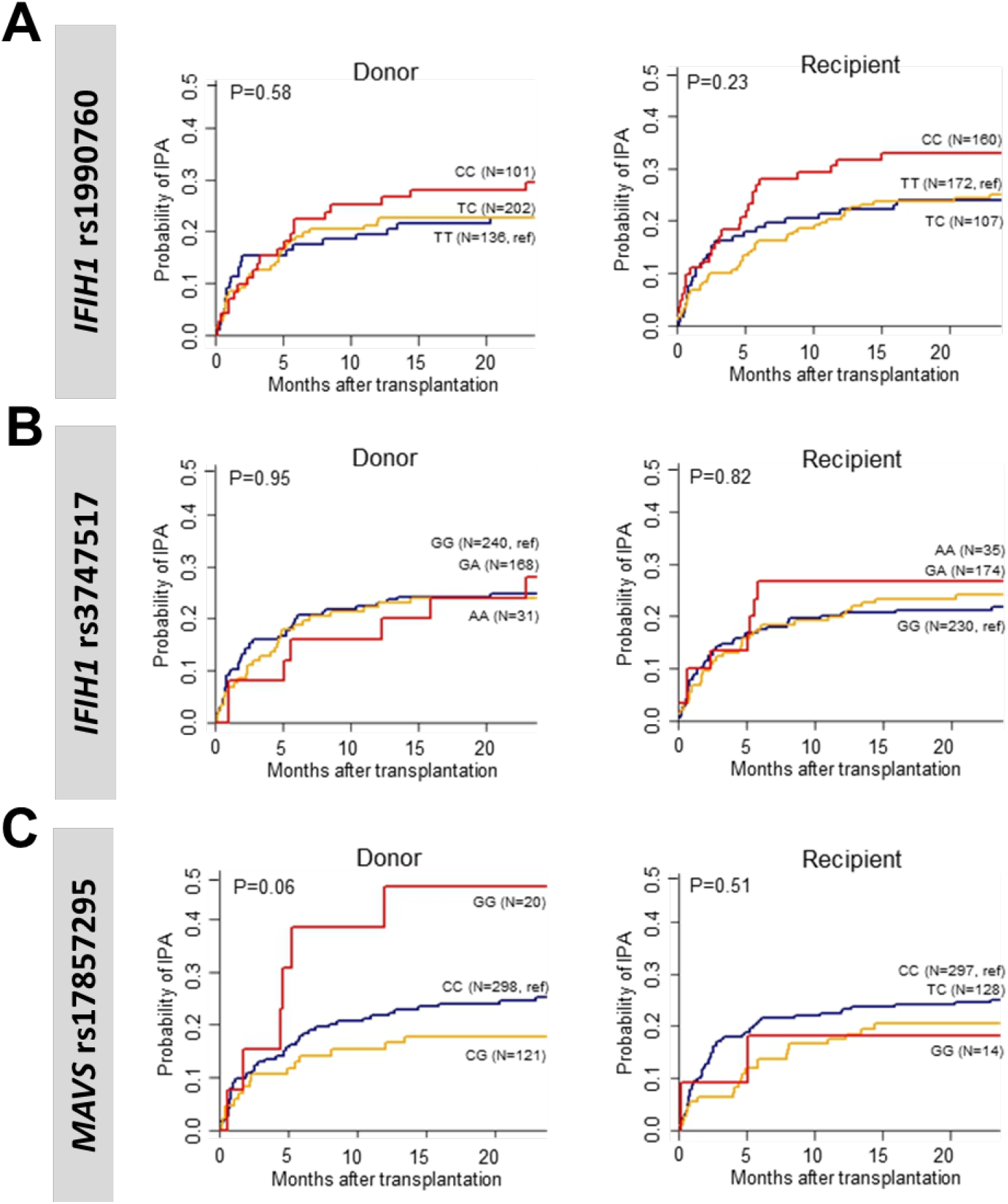
Individual genotype of *IFIH1* and *MAVS* polymorphisms in human transplant patients. Cumulative incidence analysis of invasive aspergillosis after transplantation according to donor or recipient genotypes at rs1990760 in *IFIH1* **(A)**, rs3747517 in *IFIH1* **(B)**, or rs17857295 in *MAVS* **(C)** over 24 months after HSCT. Data were analyzed by two-sided Gray’s test.

**Supplemental Figure 2.**
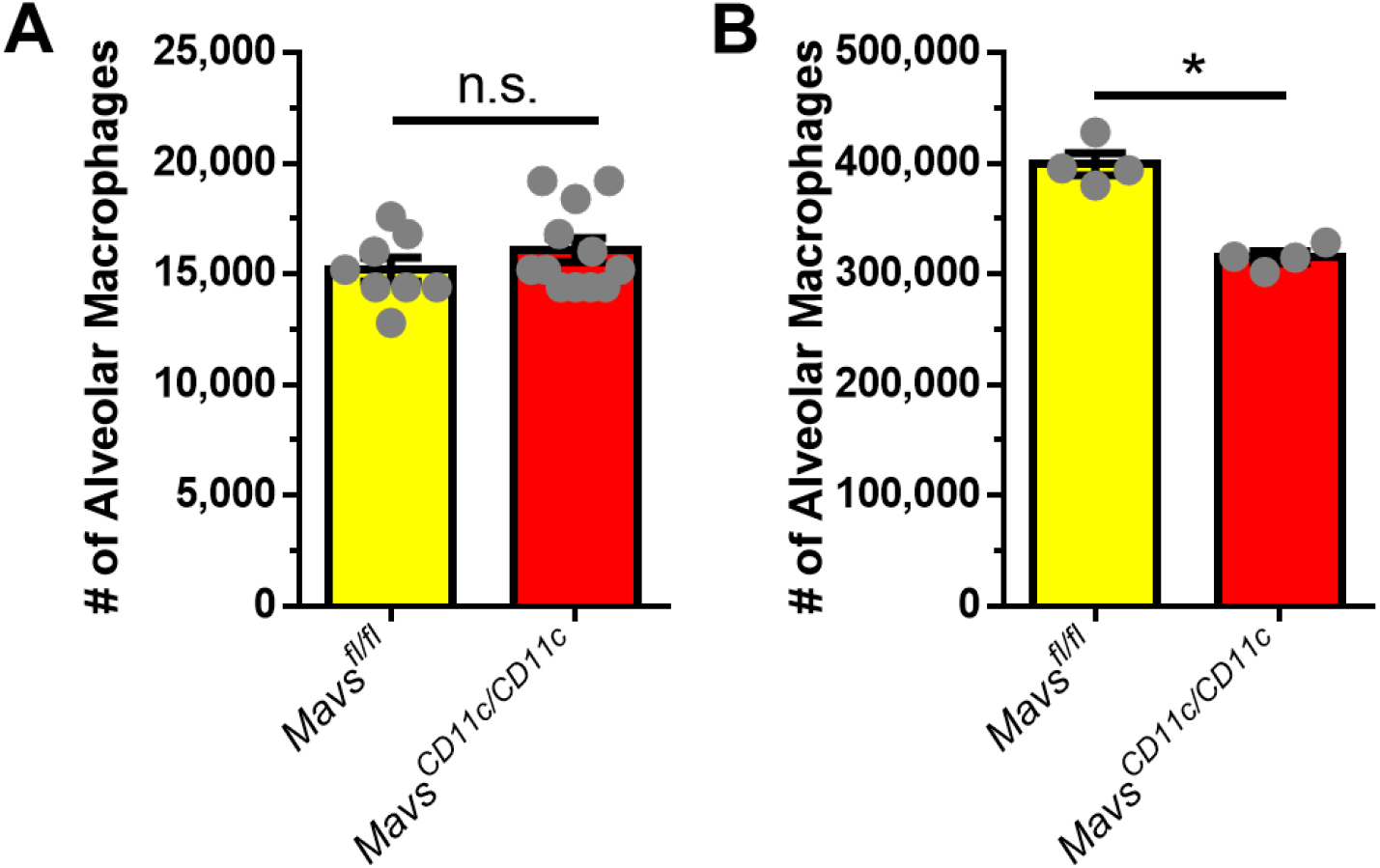
Alveolar macrophage numbers are normal in naïve mice. BAL cells from naïve *Mavs*^*<Cd11c/Cd11c>*^ *and Mavs*^*<fl/fl>*^ mice **(A)** or naïve *Mavs*^*<Cd11c/Cd11c>*^ mice given 5×10^5^ SiglecF^+^ lung cells from naïve *Mavs*^*<Cd11c/Cd11c>*^ *and Mavs*^*<fl/fl>*^ mice 24h prior **(B)** were collected in PBS/EDTA. Alveolar macrophage numbers were quantified by Cytospin analysis and differential staining. n.s. = not significant; *, P < 0.01; by Mann-Whitney U-test.

**Supplemental Figure 3.**
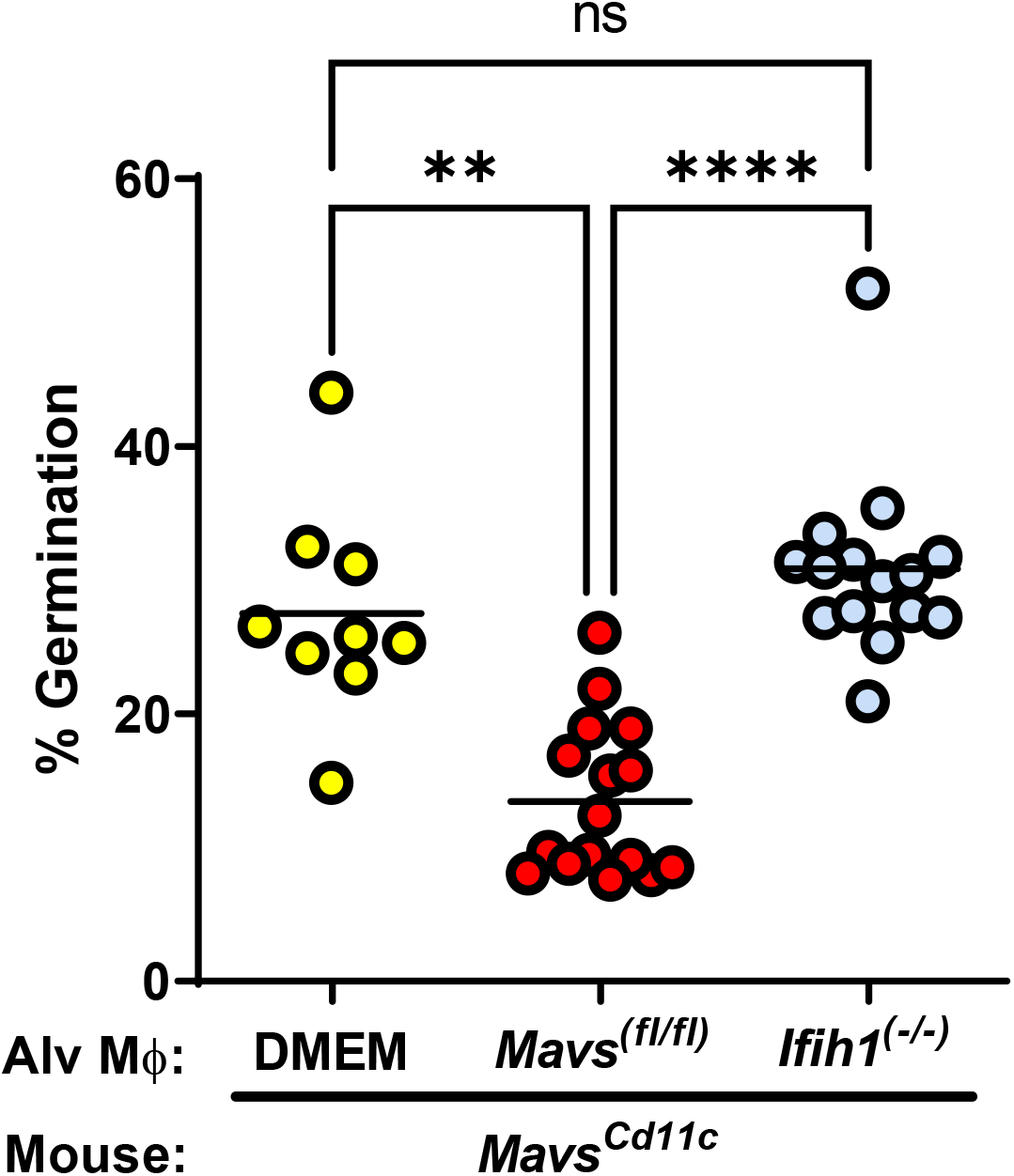
Alveolar macrophages require a Ifih1-dependent response to maintain host resistance against Aspergillus fumigatus. *Mavs*^*<Cd11c/Cd11c>*^ mice were complemented with 5×10^5^ *Ifih1*^*(-/-)*^ or *Mavs*^*(fl/fl)*^ alveolar macrophages (Siglec F^+^) given intranasally. One day later mice were challenged i.t. with 4×10^7^ resting conidia of the CEA10 isolate of *Aspergillus fumigatus*. Forty hours after *A. fumigatus* challenge mice were euthanized. Fungal germination was assessed in the lungs by GMS staining. Data are pooled from 2 independent experiments. Each dot represents an individual mouse. Statistically significant different were determined using a one-way ANOVA with a Kruskal-Wallis post-test: **p<0.01, ****p<0.0001, ns = not significant.

